# Three-Month Observational Data for the MPS IIIB Sentinel Subject Following AAV9 Mediated Gene Therapy

**DOI:** 10.64898/2026.06.01.26354386

**Authors:** Xiuwei Ma, Ruijie Gu, Wenhao Ma, Qing Xu, Rong Wang, Wenjuan Wang, Ming Liang, Xiaofang Liu, Xiao Yang, Lu Zhuang, Wendan Zhang, Xu Zeng, Juan Xu, Xiaofei Xu, Zhijie Wu, Yan Xia, Yi Liu, Jianfang Zhou, Xueyang Zhu, Hanqing Wang, Zheyue Dong, Wenyuan Yang, Yangguang Dai, Xianhui Pan, Xiu Li, Yue Wang, Xiaoyan Dong, Xiaobing Wu, Zhichun Feng

**Author notes:** These authors contributed equally to this work (Xiuwei Ma, Ruijie Gu, Wenhao Ma). Correspondence to: Zhichun Feng, MD. Seventh Medical Center of Chinese PLA General Hospital., Xiaoyan Dong, MD. Beijing Genecradle Therapeutics Inc., Beijing, China., Xiaobing Wu, MD. Beijing Genecradle Therapeutics Inc., Beijing, China.

## Abstract

**Background:** Mucopolysaccharidosis type IIIB (MPS IIIB) is a devastating neurodegenerative lysosomal storage disorder caused by alpha-N-acetylglucosaminidase (NAGLU) deficiency. There is currently no approved therapy. We report the 3-month outcomes of a novel intracerebroventricular (ICV) gene therapy in a child with MPS IIIB.

**Methods:** In an open-label, single-center, investigator-initiated trial (ChiCTR2600121466), a single dose of RDGT-101 (2.0E14vg of an AAV9 vector encoding human *NAGLU*) was administered via ICV infusion. Primary outcomes were safety and tolerability. Secondary outcomes included serum NAGLU activity, urinary heparan sulfate (HS) excretion, and neurocognitive function. Exploratory analyses included hematological parameters.

**Results:** The patient achieved serum NAGLU activity (17.06 nmol/mL/hour) approaching that of healthy controls (17.75 ± 1.37 nmol/mL/hour) by Month 3, accompanied by a 58.4% reduction in urinary HS. Clinically, previously severe hand and toe contractures resolved, allowing for full extension. Neurocognitive improvements were observed, including clear articulation, logical conversation, and sustained eye contact. Hematological analyses revealed normalized red blood cell indices and improved iron utilization. No dose-limiting toxicities, serious adverse events, or clinically significant laboratory abnormalities were observed.

**Conclusions:** A single ICV infusion of RDGT-101 was safe and well-tolerated in this patient with MPS IIIB. Early biochemical correction was accompanied by marked improvements in somatic, neurocognitive, and hematological parameters. These findings support further investigation of ICV AAV9 gene therapy for MPS IIIB.

## Introduction

Mucopolysaccharidoses (MPS) are a group of rare, progressive lysosomal storage disorders caused by deficiencies in specific enzymes required for glycosaminoglycan (GAG) catabolism [1–4]. While seven distinct clinical subtypes (MPS I–VII) are currently recognized—with the historical designation MPS V now subsumed under MPS I—inheritance patterns vary; most are autosomal recessive, whereas MPS II is X-linked [1–4]. The resulting accumulation of partially degraded GAGs induces widespread cellular dysfunction and multiorgan involvement, posing substantial clinical management challenges [1–4]. Among these, MPS IIIB is a rare autosomal recessive subtype arising from mutations in the NAGLU gene, which encodes α-N-acetylglucosaminidase [4, 5]. This enzymatic defect specifically impairs heparan sulfate (HS) degradation, leading to toxic GAG accumulation within lysosomes [5]. Unlike other MPS disorders that present with severe somatic burden, MPS IIIB exhibits a unique ’neurocentric’ phenotype [1–5]. It is characterized by relatively mild skeletal and visceral manifestations yet profound neurodegeneration, invariably leading to rapid cognitive and behavioral decline and premature death—typically in the second decade of life—despite preserved physical stature [4–6].

Reliable epidemiological data on the prevalence of MPS IIIB remain limited. Estimates from the National Organization for Rare Disorders and the National MPS Society indicate an incidence of approximately 1 in 70,000 live births for MPS III overall, with MPS IIIA and IIIB representing the most prevalent phenotypes [7,8]. Currently, there is no cure for MPS IIIB, and clinical management is restricted to palliative care [9]. Therapeutic strategies under investigation include enzyme replacement therapy (ERT), hematopoietic stem cell transplantation (HSCT), and gene therapy [4]. While regular intravenous ERT can reduce visceral HS accumulation and alleviate somatic symptoms, it fails to effectively ameliorate core neurological manifestations due to its inability to cross the blood-brain barrier [10]. Although intrathecal or intraventricular ERT delivery is under preclinical or early clinical investigation [11], ERT is typically constrained by a short half-life, necessitating frequent biweekly infusions [10,11]. This regimen incurs prohibitive costs and poses a risk of immunogenicity, which may compromise long-term efficacy [10,11].

To date, no AAV gene therapy for MPS IIIB has been approved worldwide. Preclinical studies have demonstrated that intracranial delivery of AAV-NAGLU improves motor function, auditory performance, and extends survival in murine models [4,12,13]. Clinically, ABO-101 (Abeona Therapeutics), an AAV9-based vector, received FDA Fast Track designation in 2019; however, Abeona formally discontinued its development in 2022 [14,15]. Similarly, the intracerebral AAV gene therapy program advanced by UniQure Biopharma has also been terminated [16]. Although early-phase trials have reported safety and efficacy following intracerebral administration of rAAV2/5 encoding human *NAGLU* [17], these advances have not translated into approved therapies. Consequently, there remains a critical unmet medical need for effective AAV gene therapies targeting MPS IIIB.

In this study, we report findings from a sentinel patient, enrolled in a single-arm, open-label, single-center, single-dose investigator-initiated trial (IIT) (ChiCTR2600121466). The patient received a single intracerebroventricular (ICV) infusion of RDGT-101, a next-generation AAV9-mediated gene therapy vector designed to express the human *NAGLU*, at a dose of 2.0 × 10¹⁴ vg/patient.

## Methods

### Study Design

This is an exploratory, single-arm, open-label, single-center, single-dose investigator-initiated trial (IIT) (ChiCTR2600121466). The study subjects consist of pediatric patients aged ≤12 years with MPS IIIB. A total of 6 patients are planned for enrollment, with a dose of 2.0E14 vg per patient, and a total observation period of 52 weeks. The study comprises three phases: the Screening/Baseline Period (Days -14 to -1), the Dosing Period (Day 1), and the Follow-up Period (Days 2 to Month 12). Eligible patients meeting inclusion criteria and not meeting exclusion criteria will receive the investigational treatment, followed by the follow-up period. At the end of the follow-up period, subjects, guardians, or parents will decide whether to proceed to the long-term follow-up study (Figure 1).

**Figure 1.**
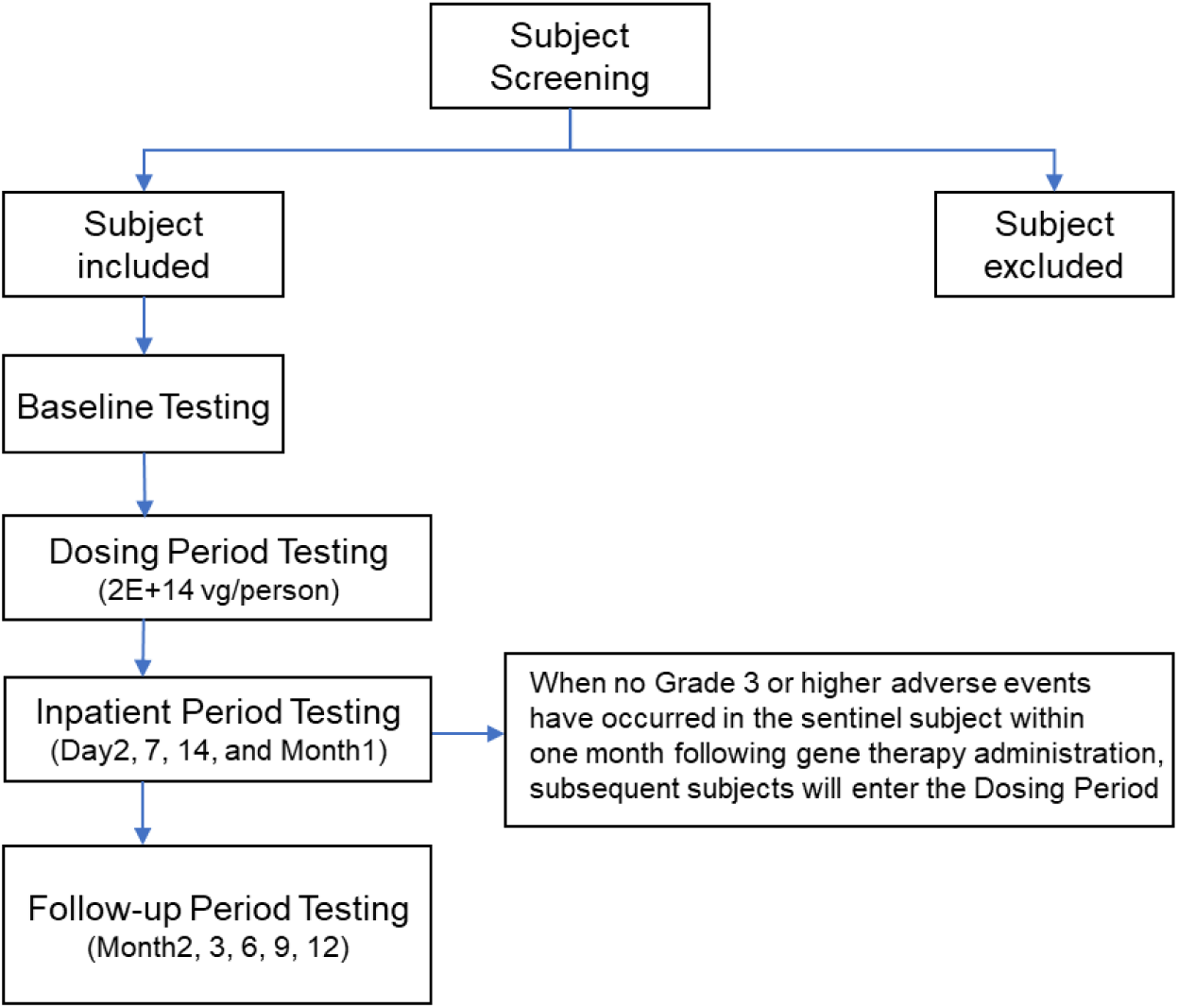
Study flowchart. A total of 6 patients are planned for enrollment in this study, with a dose of 2.0E+14 vg per patient, and a total observation period of 52 weeks. The inclusion criteria, exclusion criteria, and medical assessments at each time point illustrated in the figure are described in detail in the Methods section.

The primary endpoint was the safety profile following a single intraventricular injection of RDGT-101, assessed up to 52 weeks post-administration. Secondary endpoints encompassed efficacy, pharmacokinetics (PK), and immunogenicity, evaluated over the same 52-week period. Efficacy measures included changes from baseline in biomarkers (plasma, urinary, and cerebrospinal fluid [CSF] HS; plasma and CSF NAGLU enzyme activity; and plasma and urinary GAGs), as well as neurocognitive outcomes and quality of life (QoL), measured by Cognitive Age Equivalents, Adaptive Behavior Composite scores, and the Pediatric Quality of Life Inventory™ (PedsQL™) Generic Core Scales total score. PK analysis focused on vector genome copy numbers in whole blood, while immunogenicity was determined by serum levels of anti-AAV9 neutralizing antibodies and anti-NAGLU protein antibodies. Exploratory endpoints included the dynamics of neurological injury markers; caregiver burden assessed via the Parenting Stress Index Short Form; and histopathological evaluation of skin biopsies.

The Ethics Committee of the Seventh Medical Center of Chinese People’s Liberation Army General Hospital gave ethical approval (No. S2026-014-03) for this work. The protocol adhered to the principles of the Declaration of Helsinki, the International Council for Harmonisation Good Clinical Practice guideline, and the Good Clinical Practice guideline of the National Medical Products Administration of China. Prior to enrollment, written informed consent was obtained from all legal guardians.

For this IIT, a rigorous internal monitoring framework was established to safeguard participant safety and ensure data integrity throughout the study duration. Central to this framework were predefined protocol stopping rules, which delineated explicit criteria for implementing temporary safety pauses or full study termination in response to emerging risks. Complementing these rules, institutional internal review mechanisms were operationalized through regular safety review meetings conducted by the research team, during which cumulative safety data were systematically assessed to inform ongoing risk-benefit evaluations. Additionally, direct oversight was maintained by the principal investigator and core research team, who actively monitored participant safety, protocol adherence, and data quality in real time, thereby ensuring timely identification and management of any issues arising during the conduct of the trial.

### Participants

Participant eligibility was determined through a structured screening process guided by predefined inclusion and exclusion criteria. Inclusion required participants to be under 12 years of age, irrespective of sex, with a genetically confirmed diagnosis of MPS IIIB based on homozygous or compound heterozygous mutations in the *NAGLU* and plasma NAGLU enzyme activity at or below 10% of the established normal reference range. Exclusion criteria were comprehensive and designed to minimize confounding factors and safeguard participant welfare, encompassing advanced disease stages (e.g., severe sensory impairment, uncontrolled seizures, or loss of ambulation), evidence of symptomatic improvement, non-MPS IIIB-related CNS or behavioral disorders, prior HSCT or exposure to cellular/gene therapies, elevated anti-AAV9 neutralizing antibodies (≥1:100 titer), significant hematologic, hepatic, renal, cardiac, or metabolic abnormalities, active infections, contraindications to glucocorticoids, MRI, or intrathecal procedures, any condition potentially impairing data interpretation or safety (including immunodeficiencies, organ dysfunction, or CSF circulation disorders), positive serology for HIV, HBV, HCV, or syphilis, recent vaccination within two weeks, and any additional concerns identified by the investigator.

### Investigational Product Description

RDGT-101 Injection (hereinafter referred to as “RDGT-101”) is an investigational AAV-based therapeutic agent manufactured by Wujiahe Gene Technology Co., Ltd. and supplied by Beijing Jinlan Gene Technology Co., Ltd. RDGT-101 utilizes a recombinant adeno-associated virus serotype 9 (rAAV9) vector carrying a single-stranded transgene that encodes the human *NAGLU* under the control of a hybrid cytomegalovirus enhancer/chicken beta-actin promoter (CAGβmut). Following administration, the rAAV9 vector delivers the functional *NAGLU* and its expression cassette into the patient’s body. Sustained transgene expression restores deficient NAGLU enzyme activity, thereby reducing the abnormal accumulation of HS and re-establishing normal HS metabolic homeostasis.

### Route of Administration of Gene Therapy

The procedure will be performed under general anesthesia in the operating room. The patient will be placed in the supine position and prepared in a sterile fashion. A right-sided frontal burr hole will be created at a point 2 cm posterior to the hairline and 2 cm lateral to the midline. Using a ventricular puncture needle advanced parallel to the sagittal plane and perpendicular to the line connecting the external auditory canals (interaural line), the needle will be inserted to a depth of 6 cm. The stylet will then be removed, and clear CSF will be confirmed. A 4 mL dose of RDGT-101 will be slowly injected into the lateral ventricle. Following completion of the infusion, the needle will be withdrawn, local pressure applied, and a sterile dressing placed over the site. Vital signs will be continuously monitored throughout the procedure and during the immediate postoperative period. Postoperative management will include anti-infective prophylaxis, blood pressure control, acid suppression therapy, and close observation for any clinical changes

### Subject Observation and Assessments

This single-dose clinical study plans to enroll a total of 6 subjects. A staggered enrollment design will be implemented, wherein the first subject will be observed for at least 4 weeks post-dosing. Investigators must conduct a comprehensive safety analysis based on data obtained from dosing through the first 4 weeks. If no Grade 3 or higher adverse events (per CTCAE v5.0) occur, enrollment and dosing of the second subject will be permitted. Subsequent subjects will follow the same procedure until all enrolled patients have received the investigational product. In the event of a Grade 3 or higher adverse event, investigators must thoroughly assess the relationship to the study drug before deciding whether to suspend enrollment. All subjects who receive the investigational product will undergo continuous safety assessments and efficacy tracking for 52 weeks.

Safety evaluations included vital signs, physical examinations, 12-lead electrocardiograms (ECGs), Doppler echocardiography, clinical laboratory tests, etiology screening, and adverse event (AE) monitoring.

Vital sign parameters comprised blood pressure, heart rate, respiratory rate, and body temperature. The timing of assessments is detailed in the “Study Timeline”. Measurements obtained prior to dosing on Day 1 (at the time point closest to administration) were designated as the baseline for safety analysis. Any clinically significant abnormalities compared to baseline were reported as adverse events.

Comprehensive physical examinations covered the following systems: skin and mucosa, lymph nodes, head, neck, chest, abdomen (including abdominal circumference measurement), musculoskeletal system, muscle tone, extremities, and the nervous system.

Immunogenicity Assessments: Anti-AAV9 binding antibody and anti-NAGLU protein antibody levels at pre-dosing (screening/baseline), Month 3, Month 6, Month 12, and at early withdrawal. Anti-AAV9 neutralizing antibody levels at pre-dosing (screening/baseline), Month 1, Month 2, Month 3, Month 6, Month 9, Month 12, and at early withdrawal. Peripheral blood mononuclear cell (PBMC) samples were collected for analysis two weeks after discontinuation of prednisolone. Additionally, if an adverse event occurred (e.g., significant elevation of AST or ALT) and the investigator deemed it necessary, cellular immune response assessments could be performed outside the scheduled time points (at the time of the event and during recovery) to evaluate a potential correlation between the event and the immune response. Details regarding the immunological evaluation methods can be found in our previous report [18].

Efficacy Evaluation Parameters: 1. Levels of the substrate HS in CSF were measured at pre-dosing (screening/baseline) and at 6 and 12 months post-dosing. HS levels in plasma and urine were measured at pre-dosing (screening/baseline) and at 2 weeks, and 1, 2, 3, 6, 9, and 12 months post-dosing; 2. NAGLU enzyme activity in CSF was measured at pre-dosing (screening/baseline) and at 6 and 12 months post-dosing. NAGLU enzyme activity in plasma was measured at pre-dosing (screening/baseline) and at 2 weeks, and 1, 2, 3, 6, 9, and 12 months post-dosing; 3. Plasma and urinary GAG levels were measured at pre-dosing (screening/baseline) and at 2 weeks, and 1, 2, 3, 6, 9, and 12 months post-dosing; 4. Brain volume, cerebral structure, and lesions (e.g., brain atrophy, white matter lesions, and other pathologies) were assessed via MRI at pre-dosing (screening/baseline) and at 6 and 12 months post-dosing; 5. Liver and spleen volumes were measured via abdominal ultrasound at pre-dosing (screening/baseline) and at 3, 6, and 12 months post-dosing; 6. Changes in Cognitive Age Equivalents and Developmental Quotient (DQ) from baseline were evaluated using the Pediatric Developmental Assessment Scale at 1, 2, 3, 6, and 12 months post-dosing. Results were compared with natural history data; 7. Changes in Adaptive Behavior Composite scores (Age Equivalent Scores) from baseline were evaluated using the Infant-Junior High School Student Adaptive Behavior Scale at 1, 2, 3, 6, and 12 months post-dosing. Results were compared with natural history data; 8. Changes in the total score of the Pediatric Quality of Life Inventory™ (PedsQL™) Generic Core Scales from baseline were evaluated at 1, 2, 3, 6, and 12 months post-dosing; and 9. Blood samples were collected for PK analysis at pre-dosing (screening/baseline), Day 1, Day 2, Day 7, Day 14, Month 1, Month 2, Month 3, Month 6, Month 9, Month 12, and at early withdrawal.

### Measurement of Vector Genomes in Blood

Blood samples for vector genome detection were collected according to the protocol outlined [18]. Genomic DNA was extracted from 100 μL of EDTA-K2 anticoagulated whole blood using the QIAamp DNA Mini Kit (QIAGEN, Hilden, Germany). Vector genome copy numbers were quantified using a digital droplet PCR (ddPCR) assay with primers and probes specific to the rAAV9-coGAA vector. The sequences used were as follows: forward primer 5’-GGCACCTGGTCGTGGAACTC-3’; reverse primer 5’-CCAGCCTCTGATCGGCAAGG-3’; and probe [FAM] 5’-TGGCCAGGCTCCACCGCCTT-3’ [TAMRA]. All oligonucleotides were synthesized by Sangon Biotech (Shanghai, China). Each sample was tested in duplicate, and results are expressed as genome copies per milliliter (copies/mL) of blood. The lower limit of detection was determined to be 2.5 × 10⁴ copies/mL.

### NAGLU Enzyme Activity Assay

The enzyme activity was measured with a fluorogenic substrate 4-methylumbelliferyl-2-acetamido-2-deoxy-a-o-glucopyranoside (4MU α GlcNAc) as previously reported [19]. Briefly, samples are incubated with the artificial substrate 4-MU α GlcNAc in an acidic acetate buffer (pH 3.7) at 37°C for about 17hs. The enzymatic reaction is terminated by adding an alkaline glycine-NaOH buffer (pH ∼10.7) to allow the fluorescent product, 4-methylumbelliferone, to be measured. Fluorescence intensity is recorded using a plate reader (excitation ∼360 nm, emission ∼460 nm), and enzyme activity is quantified by comparing the signal to a standard curve generated from known concentrations of 4-methylumbelliferone, then normalized to sample volume.

### Urinary HS measurement

Urinary HS levels were determined using a targeted Liquid Chromatography–Tandem Mass Spectrometry (LC–MS/MS) assay optimized for MPS III disease monitoring [20]. First morning urine specimens were collected, centrifuged to remove cellular debris, and stored at –80 °C until analysis. HS glycosaminoglycan chains were enzymatically digested into disaccharides using a combination of heparin lyases I, II, and III under controlled temperature and pH conditions. Following enzymatic digestion, samples were purified by solid-phase extraction, concentrated under nitrogen, and reconstituted in mobile phase-compatible solvent. Chromatographic separation was achieved on a hydrophilic interaction liquid chromatography (HILIC) column with an ammonium acetate–acetonitrile gradient. Detection was performed in negative-ion electrospray ionization mode using multiple reaction monitoring (MRM) transitions specific for HS-derived disaccharides, including those with characteristic sulfation patterns associated with MPS III pathology. Stable isotope-labeled internal standards were used for each targeted disaccharide to ensure quantitative accuracy. Calibration curves were prepared from synthetic HS disaccharide standards. To correct for variations in urine concentration, HS disaccharide concentrations were normalized to urinary creatinine. Creatinine levels were measured in the same urine samples using a validated colorimetric Jaffé method or enzymatic assay, and HS values were expressed as nanomoles of HS disaccharide per millimole of creatinine (nmol/mmol Cr). This normalization minimizes the impact of hydration status and diurnal variation on biomarker quantification.

## Results

### Clinical History and Baseline Characteristics of the Sentinel Case

This male patient was born via full-term spontaneous vaginal delivery (G1P1), with a birth weight of 3.7 kg. Developmental delays were first noted at early childhood (*to avoid specifying age for ethical reasons*), manifesting as delayed language acquisition, inability to integrate into class activities, and short stature. With increasing age, although capable of expressing basic needs, the patient exhibited unclear articulation, occasional tangential responses, poor eye contact, and limited interest in play with predominantly sedentary behavior. He also suffered from poor appetite and frequent gagging during meals. Physical examination revealed coarse facies, coarse skin, hand contractures limiting extension, and toe clawing.

Enzymatic analysis confirmed MPS IIIB, revealing significantly deficient NAGLU activity (2.2 nmol/g/h). Genetic testing identified compound heterozygous variants: a maternal c.1384G>C (p.Gly462Arg) and a paternal c.607C>T (p.Arg203)*.

Baseline Clinical Profile. Anthropometric measurements showed a height of 118 cm, weight of 26 kg, and a BMI of 18.7 kg/m². Cardiac evaluation indicated posterior mitral leaflet prolapse with moderate mitral regurgitation (MR), although left ventricular systolic function was preserved. Brain MRI demonstrated cerebral atrophy and minimal white matter lesions. Quality of life assessment using the PedsQL 4.0 questionnaire indicated marked impairments, particularly in social functioning (5.0) and school functioning (35.0), alongside reduced physical functioning (18.75) and emotional functioning (75.0). Palpable hepatosplenomegaly was noted, consistent with abdominal circumference measurements showing 61 cm and 59 cm at the second and third months post-treatment, respectively.

### Primary Outcome Measures: Safety Evaluation per CTCAE v5.0

Safety observations during the first three months post-RDGT-101 infusion were favorable. Of the 14 adverse events (AEs) listed in Table 2, only two mild (Grade 1) events were recorded: one episode of transient hypokalemia and one episode of transient pericardial effusion. Importantly, none of these AEs were assessed as serious or considered possibly related to the investigational product. No Grade ≥2 toxicities, infusion-related reactions, or clinically significant laboratory abnormalities were observed during this period, supporting an early safety profile characterized by good tolerability.

**Table 1.** Baseline characteristics of the sentinel case.

| **Table 1. Baseline characteristics of the sentinel case.** |  |
| --- | --- |
| Sex | Male |
| Birth | G1P1, full-term vaginal delivery, birth weight 3.7 kg |
| ***Characteristics at diagnosis*** |  |
| Age (years) | 5 to 10 |
| Serum enzymatic activity | 2.2 nmol/g/h |
| Total urinary GAG concentration | Not Tested |
| NAGLU mutations |  |
| Maternal-origin | c.1384G>C, p.G462R, Variant of uncertain significance |
| Paternal-origin | c.607C>T, p.R203X |
| ***Pre-enrollment history*** |  |
| The parents noted delays in language development, difficulty participating in class activities, and short stature (years) | 5 to 10 |
| Able to communicate simply and express needs, but with unclear articulation and occasional irrelevant answers; hyperactive with inability to sit still; unsteady gait (years) | 5 to 10 |
| Epilepsy | No seizures |
| ***Baseline tests*** |  |
| Age (years) | 5 to 10 |
| Coarse features | Yes |
| Height (cm) | 118 |
| Weight (kg) | 26 |
| BMI (kg/m<sup>2</sup>) | 18.7 |
| Abdominal circumference (cm) | 61 (month 2) and 59 (month 3) post-treatment |
| Head circumference (cm) | Not Tested |
| Hepatomegaly | Yes |
| Splenomegaly | Yes |
| Echocardiography | Posterior mitral leaflet prolapse with moderate MR; LV systolic function normal |
| ECG | Sinus arrhythmia, essentially normal ECG |
| Chest X-ray | Increased lung markings bilaterally; no rib abnormalities detected |
| Brain MRI | Cerebral atrophy and minimal white matter lesions |
| HS concentration in CSF (ng/ml) | Not Tested |
| ***PedsQL 4.0 Generic Core Scales*** |  |
| Physical functioning | 18.75 |
| Emotional functioning | 75 |
| Social functioning | 5 |
| Learning | 35 |
| BMI, body mass index; CSF, cerebrospinal fluid; ECG, electrocardiogram; GAG, glycosaminoglycan; HS, heparan sulphate; NAGLU, N-acetylglucosaminidase. |  |

**Table 2.** Adverse events during the 3-month observation period following RDGT-101 treatment.

| | Number of events | Number of events (grade $\geq 3$) | Possibly drug-related adverse events | Serious adverse events |
| --- | --- | --- | --- | --- |
| Fever | 0 | 0 | 0 | 0 |
| Upper respiratory tract infection | 0 | 0 | 0 | 0 |
| Pneumonia and bronchitis | 0 | 0 | 0 | 0 |
| Influenza | 0 | 0 | 0 | 0 |
| Cough | 0 | 0 | 0 | 0 |
| ALT increased | 0 | 0 | 0 | 0 |
| AST increased | 0 | 0 | 0 | 0 |
| ALP increased | 0 | 0 | 0 | 0 |
| Hypertriglyceridemia | 0 | 0 | 0 | 0 |
| Hypercholesterolemia | 0 | 0 | 0 | 0 |
| Constipation | 0 | 0 | 0 | 0 |
| Diarrhea | 0 | 0 | 0 | 0 |
| Functional gastrointestinal disorder | 0 | 0 | 0 | 0 |
| Eczema | 0 | 0 | 0 | 0 |
| Aphtha | 0 | 0 | 0 | 0 |
| Anaemia | 0 | 0 | 0 | 0 |
| Hypoxemia | 0 | 0 | 0 | 0 |
| Hypercapnia | 0 | 0 | 0 | 0 |
| Acidosis | 0 | 0 | 0 | 0 |
| Urinary tract infection | 0 | 0 | 0 | 0 |
| Right leg movement restricted | 0 | 0 | 0 | 0 |
| Hypokalemia | 1 | 0 | 0 | 0 |
| Pericardial effusion | 1 | 0 | 0 | 0 |
| Total | 2 | 0 | 0 | 0 |

According to the Common Terminology Criteria for Adverse Events 5.0 criteria, Grade 1: Mild; asymptomatic or mild symptoms; clinical or diagnostic observations only; intervention not indicated. Grade 2: Moderate; minimal, local or noninvasive intervention indicated; limiting age-appropriate instrumental activities of daily living. Grade 3: Severe or medically significant but not immediately life-threatening; hospitalization or prolongation of hospitalization indicated; disabling; limiting self care activities of daily living. Grade 4: Life-threatening consequences; urgent intervention indicated. Grade 5: Death related to adverse event.

### Primary Outcome Measure: Safety Evaluation Based on Biochemical Markers

As shown in Figure 2, longitudinal assessment of serum biomarkers demonstrated that hepatic function, as indicated by alanine aminotransferase (Panel A), aspartate aminotransferase (Panel B), and alkaline phosphatase (Panel C), remained consistently within the normal reference ranges throughout the 3-month follow-up period, suggesting the absence of hepatotoxicity. Renal function, assessed by urea (Panel D) and creatinine (Panel E) levels, also remained stable within normal limits, indicating no evidence of nephrotoxicity. Furthermore, markers of cellular damage and cardiac stress, including cardiac troponin I (Panel F), creatine kinase (Panel G), and lactate dehydrogenase (Panel H), showed no significant elevations above the normal reference values. This indicates the absence of cardiac injury, muscle damage, or cell damage or tissue necrosis. In conclusion, the gene therapy was well-tolerated, with no clinically significant abnormalities observed in liver, renal, or cardiac biochemical parameters, supporting its favorable safety profile in MPS IIIB patient.

**Figure 2.**
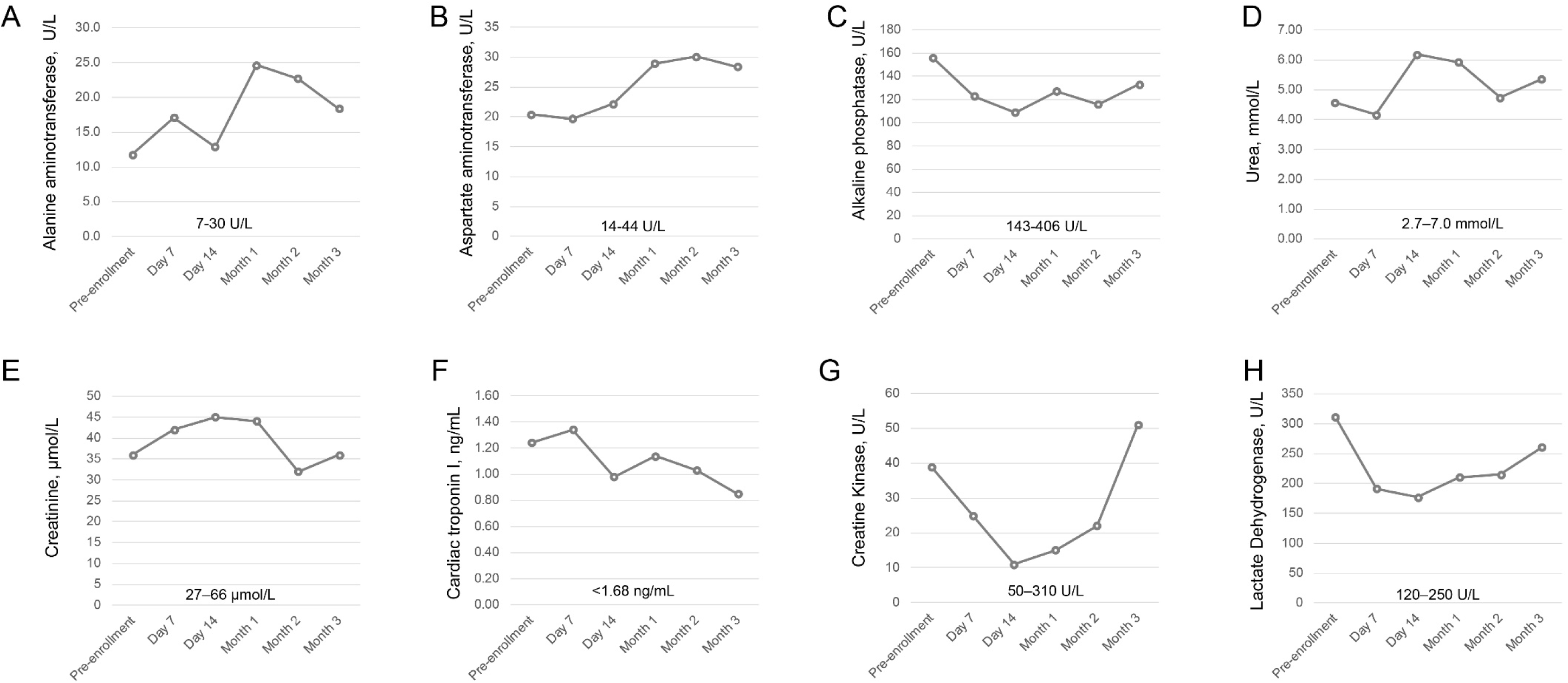
Clinical biochemistry markers indicating organ function before and after gene therapy. Longitudinal changes in serum biochemical markers were monitored to assess potential organ toxicity in MPS IIIB patient following gene therapy. Liver function: (A) Alanine aminotransferase, (B) Aspartate aminotransferase and (C) Alkaline phosphatase levels. Renal function: (D) Blood urea and (E) Serum creatinine levels. Cardiac and Muscle function: (F) Cardiac troponin I, (G) Creatine Kinase (CK) and (H) Lactate Dehydrogenase (LDH) levels.

### Secondary Outcome Measures

The kinetics of RDGT-101 vector genomes in the peripheral blood were monitored over time. At Day 2 post-administration, the vector DNA copy number peaked at 9.78 × 10⁸ ± 1.45 × 10⁸ copies/mL. This was followed by a rapid clearance phase, with levels dropping to 6.12 × 10⁶ ± 3.06 × 10⁵ copies/mL by Day 7 and further declining to 1.46 × 10⁶ ± 2.26 × 10⁴ copies/mL by Month 3, indicating a sustained reduction of vector DNA in circulation (Figure 3A).

**Figure 3.**
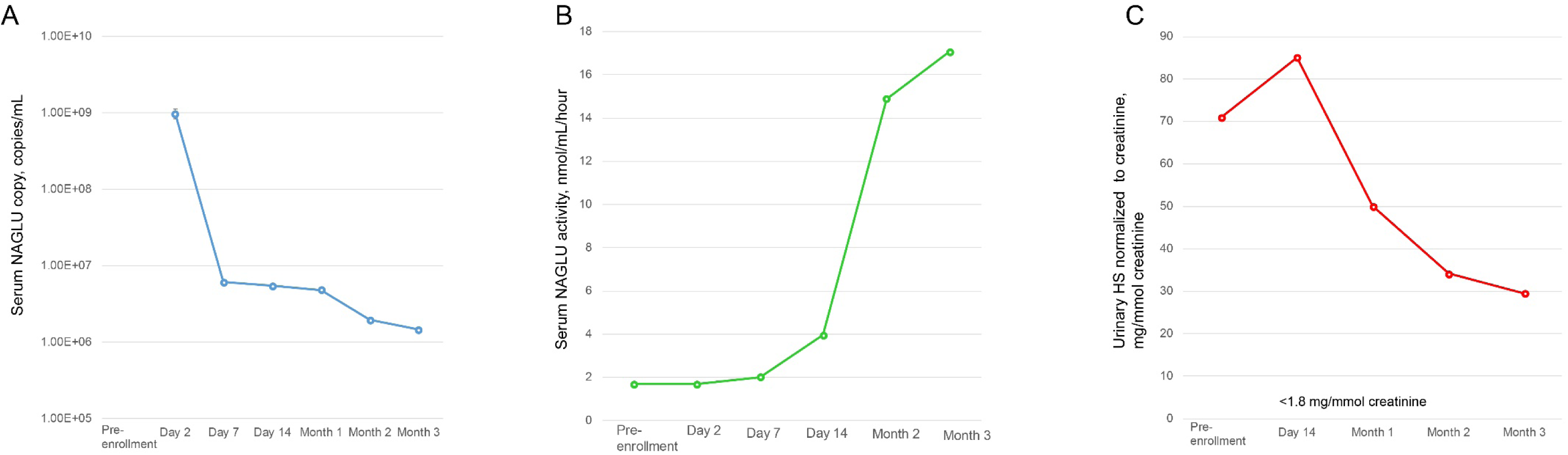
Monitoring of serum NAGLU DNA copies, NAGLU enzyme activity, and urinary HS concentration. A. Serum NAGLU DNA copy number after gene therapy. B. Changes in serum NAGLU enzyme activity before and after gene therapy. C. Changes in urinary HS concentration before and after gene therapy. HS values were expressed as nanomoles of HS disaccharide per millimole of creatinine.

In contrast to the dynamic trend of the vector genome, during the pre-enrollment period and at Days 2 and 7 post-treatment, NAGLU levels remained low (range: 1.68–2.02 nmol/mL/hour). A notable increase was observed by Day 14, with activity rising to 3.95 nmol/mL/hour. The activity continued to climb sharply, reaching 14.89 nmol/mL/hour at Month 2 and further increasing to 17.06 nmol/mL/hour by Month 3. This value approached the mean level of healthy controls (17.75 ± 1.37 nmol/mL/hour), indicating robust and sustained NAGLU expression (Figure 3B).

Urinary HS concentration normalized to creatinine showed dynamic changes. At pre-enrollment, the concentration was measured at 70.92 mg/mmol creatinine. It increased to a peak of 85.08 mg/mmol creatinine by Day 14 post-treatment. Following this peak, a gradual decline was noted, with levels decreasing to 50.07 mg/mmol creatinine by Month 1 and further reducing to 29.49 mg/mmol creatinine by Month 3, suggesting an initial therapeutic effect followed by metabolic stabilization (Figure 3C).

### Exploratory Analysis of Hematological Parameters

Three months post-treatment, serum NAGLU activity normalized (approaching control levels), and urinary HS excretion was reduced by 58.4%. Clinically, the patient exhibited marked improvements: appetite normalized, skin texture became smooth, and hand and toe contractures resolved, allowing for full extension. Notably, neurocognitive gains were observed, characterized by increased playfulness, clear articulation, logical conversational skills, and sustained eye contact.

To investigate whether these phenotypic improvements correlated with systemic physiological enhancements in growth, nutrition, or metabolic mobilization, we analyzed longitudinal hematological data (Figure 4, Table 3).

**Figure 4.**
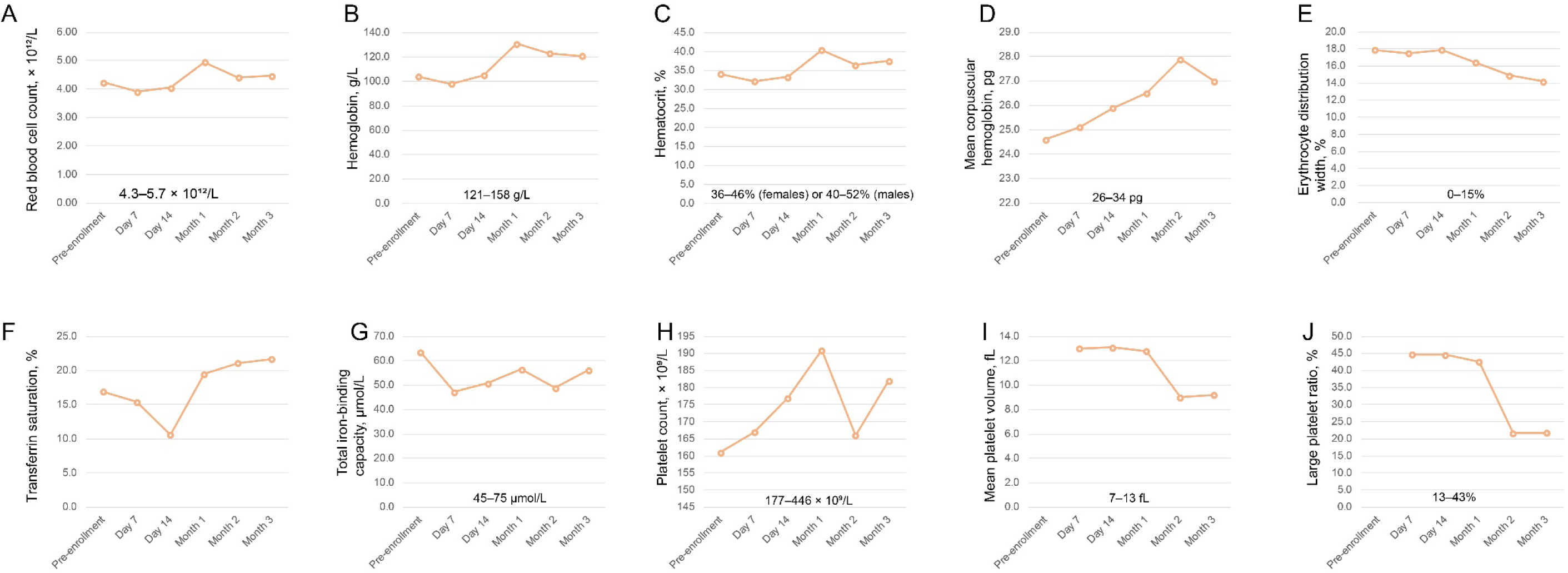
Longitudinal changes in hematological parameters during and after treatment. (A) Red blood cell count, (B) Hemoglobin, (C) Hematocrit, (D) Mean corpuscular hemoglobin, (E) Erythrocyte distribution width, (F) Transferrin saturation, (G) Total iron binding capacity, (H) Platelet count, (I) Mean platelet volume, (J) Large platelet ratio. Reference ranges are indicated below each panel.

**Table 3.**
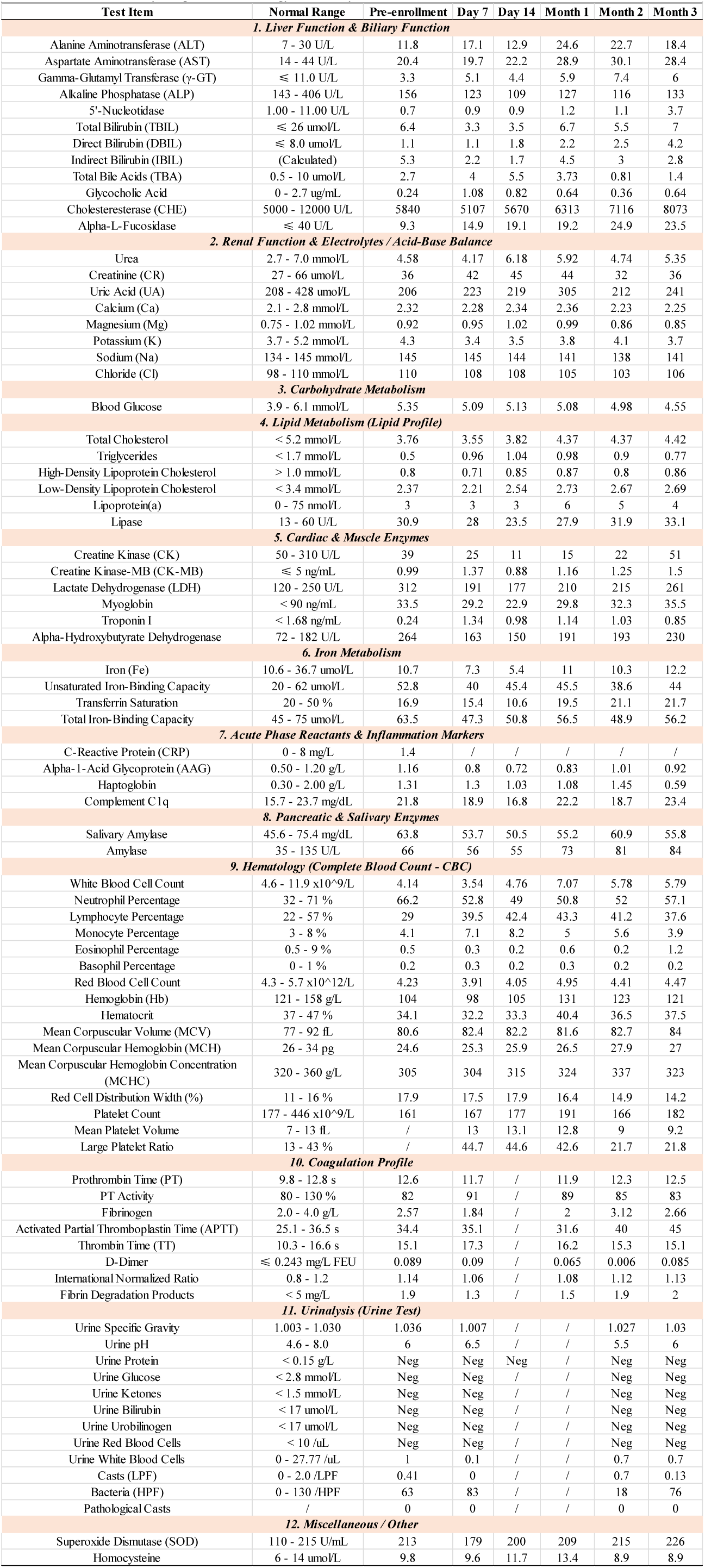
Clinical biochemistry, coagulation, hematology, and urinalysis.

Erythropoiesis and Nutritional Status (Panels A–D, Table 3): Red blood cell (RBC) indices demonstrated the most prominent changes. RBC count (Panel A) and Hemoglobin (Hb, Panel B) showed an overall upward trend and reaching the normal reference range. Concurrently, Mean Corpuscular Hemoglobin (MCH, Panel D) increased from ∼24.6 pg to ∼27.9 pg. These trends suggest enhanced erythropoietic activity and improved nutritional status following the alleviation of metabolic burden.

Iron Metabolism Dynamics (Panels F–G): Transferrin Saturation (Panel F) increased in an undulating manner from ∼16.9% to ∼21.7%, while Total Iron-Binding Capacity (TIBC, Panel G) demonstrated fluctuating stability. This pattern suggests improved iron utilization and a resolution of functional iron deficiency.

Platelet Indices (Panels H–J): Platelet Count (Panel H) remained within the normal range. Mean Platelet Volume (MPV, Panel I) and the Large Platelet Ratio (Panel J) exhibited a decreasing trend. While the clinical significance requires further validation, these results suggest improved platelet maturity and the restoration of the hematopoietic microenvironment.

Taken together, although limited to a single-patient case study, the above hematological shifts suggest an enhanced capacity for growth, tissue repair, and nutritional mobilization, correlating with observed gains in motor function and cognitive-behavioral regulation.

## Discussion

In this sentinel case report, we present the 3-month safety and efficacy outcomes of a single child with MPS IIIB treated with RDGT-101, a novel AAV9-based gene therapy vector administered via a single ICV infusion.

The primary safety findings over the first 3 months were highly encouraging. The absence of dose-limiting toxicities, serious adverse events (SAEs), or clinically laboratory abnormalities related to hepatic, renal, or cardiac function aligns with the established safety profile of AAV-mediated gene therapies delivered centrally. Notably, only two mild, transient AEs (hypokalemia and pericardial effusion) were recorded, neither of which was attributed to the investigational product. This favorable early safety profile supports the feasibility of ICV delivery for AAV9 vectors in MPS IIIB, a critical consideration given the historical challenges and safety concerns associated with high-dose systemic AAV administrations and previous intracranial gene therapy trials in other lysosomal storage disorders [17,21].

The kinetics of RDGT-101 vector genomes in peripheral blood demonstrated a rapid peak at Day 2 post-infusion followed by a steady clearance, a pattern consistent with the expected distribution and transient presence of AAV vectors in circulation following central administration. More importantly, we observed a robust and sustained restoration of serum NAGLU enzyme activity, which approached the mean level of healthy controls by Month 3. This systemic enzyme expression, likely derived from transduction of periventricular structures and subsequent enzyme secretion or cross-correction, was associated with a 58.4% reduction in urinary HS excretion (normalized to creatinine) from baseline to month 3. While the initial rise in urinary HS at Day 14 is intriguing, it may reflect an early mobilization of stored substrates from tissues as lysosomal function begins to normalize, followed by enhanced clearance and metabolic stabilization. These biochemical corrections are pivotal, as they signify the partial reversal of the primary metabolic defect responsible for cellular toxicity in MPS IIIB [22].

Clinically, the patient exhibited marked and rapid improvements across multiple domains by 3 months post-treatment. The resolution of coarse facial features and skin texture, the full extension of previously contracted hands and toes, and the normalization of appetite and gagging reflex suggest a profound impact on somatic manifestations, which are typically considered mild but discernible in MPS IIIB [23]. Crucially, the neurocognitive and behavioral gains—including clearer articulation, logical conversational skills, sustained eye contact, increased playfulness, and mischievousness—represent a significant departure from the natural history of MPS IIIB, which is characterized by relentless developmental regression, behavioral deterioration, and loss of acquired skills [4–9]. These observations align with and extend the findings from previous intracerebral AAV2/5-NAGLU trials [17–21], which also reported stabilization or improvement in neurocognitive function, albeit with different vectors, routes, and immunosuppressive regimens.

A novel aspect of our findings is the detailed longitudinal analysis of hematological parameters, which revealed concurrent improvements in erythrocyte indices (RBC count, Hb, MCH), optimized iron metabolism (increased TSAT, stabilized TIBC), and platelet maturation (decreasing MPV and P-LCR within normal ranges). While hematological abnormalities are not a primary diagnostic feature of MPS IIIB [4–9], these shifts suggest that the systemic metabolic correction achieved via ICV gene therapy may alleviate secondary stressors such as chronic inflammation, suboptimal nutrient utilization, or mild marrow microenvironment dysfunction associated with long-standing lysosomal storage [4–9]. The convergence of these hematological improvements with gains in growth, tissue repair capacity, and cognitive-behavioral regulation provides preliminary evidence that effective CNS-directed gene therapy for MPS IIIB may confer broader systemic benefits beyond the classically defined neurocentric and visceral burdens. This observation warrants further investigation in larger cohorts with extended follow-up.

Several limitations of this study must be acknowledged. Foremost, this is a single-patient case report with a limited 3-month follow-up period. While the observed changes are clinically meaningful and biologically plausible, they cannot be definitively attributed to the gene therapy alone without a control comparison or longer-term data to rule out placebo effects, natural variability, or concomitant interventions (though none were reported). The young age at treatment is noteworthy; while patients in this age range retain some plasticity, earlier intervention (prior to significant neuronal loss) is generally hypothesized to yield superior neurocognitive outcomes. Additionally, the assessment of neurocognitive function relied on parent-reported observations, rather than comprehensive, blinded neuropsychological testing batteries employed in larger trials. Finally, the biodistribution and persistence of AAV9-NAGLU transduction within the human brain, particularly in deeper cortical and subcortical regions critical for cognition, remain to be fully elucidated through advanced imaging.

In conclusion, this first-in-human (or among the first) report of RDGT-101, an ICV-administered AAV9-NAGLU gene therapy for MPS IIIB, demonstrates a favorable early safety profile and provides preliminary evidence of biochemical correction, somatic symptom resolution, and notable neurocognitive/behavioral improvements at 3 months post-treatment. The concurrent normalization of hematological parameters offers an intriguing exploratory signal of systemic metabolic benefit. These findings support the continued investigation of ICV AAV9 gene therapy for MPS IIIB and highlight the potential for early intervention to alter the devastating disease trajectory. Further enrollment in the ongoing trial (ChiCTR2600121466), longer-term follow-up, and the incorporation of more sensitive neurocognitive assessments will be essential to confirm these initial promising results and to define the optimal therapeutic window for MPS IIIB gene therapy.

## Data Availability

All data produced in the present study are available upon reasonable request to the authors

## Acknowledgement

We thank the children and their families for their support of our study. We also acknowledge the support provided by the ’Cradle Program’ at the Ruicy Institute during the preclinical stage of this project.

## References

1. Sahin O, Thompson HP, Goodman GW, Li J, Urayama A. Mucopolysaccharidoses and the blood-brain barrier. Fluids Barriers CNS. 2022 Sep 19;19(1):76. doi: 10.1186/s12987-022-00373-5.

2. Lipiński P, Różdżyńska-Świątkowska A, Wiśniewska K, et al. Mucopolysaccharidoses-What Clinicians Need to Know: A Clinical, Biochemical, and Molecular Overview. Biomolecules. 2025;15(10):1448. Published 2025 Oct 14. doi:10.3390/biom15101448

3. The Metabolic and Molecular Bases of Inherited Disease (Scriver, C. R., Beaudet, A. L., Sly, W. S., Valle, D., Childs, B., Kinzler, K. W., and Vogelstein, B., eds., 8th ed., McGraw-Hill, New-York, 2001, 7012 p., $550.00). Biochemistry (Moscow) 67, 611–612 (2002). 10.1023/A:1017418800320

4. Wagner VF, Northrup H. Mucopolysaccharidosis Type III. 2019 Sep 19. In: Adam MP, Bick S, Mirzaa GM, et al., editors. GeneReviews® [Internet]. Seattle (WA): University of Washington, Seattle; 1993-2026. Available from: https://www.ncbi.nlm.nih.gov/books/NBK546574/.

5. Beesley CE, Jackson M, Young EP, Vellodi A, Winchester BG. Molecular defects in Sanfilippo syndrome type B (mucopolysaccharidosis IIIB). J Inherit Metab Dis. 2005;28(5):759–767. doi:10.1007/s10545-005-0093-y

6. Spahiu L, Behluli E, Peterlin B, Nefic H, Hadziselimovic R, Liehr T, Temaj G. Mucopolysaccharidosis III: Molecular basis and treatment. Pediatr Endocrinol Diabetes Metab. 2021;27(3):201–208. doi: 10.5114/pedm.2021.109270.

7. National Organization for Rare Disorders. https://rarediseases.org/rare-diseases/. Accessed on May 16, 2026

8. The National MPS Society. https://mpssociety.org/en/. Accessed on May 16, 2026

9. Spahiu L, Behluli E, Peterlin B, et al. Mucopolysaccharidosis III: Molecular basis and treatment. Mukopolisacharydoza III: podstawy molekularne i leczenie. Pediatr Endocrinol Diabetes Metab. 2021;27(3):201–208. doi:10.5114/pedm.2021.109270

10. Taylor M, Khan S, Stapleton M, et al. Hematopoietic Stem Cell Transplantation for Mucopolysaccharidoses: Past, Present, and Future. Biol Blood Marrow Transplant. 2019;25(7):e226–e246. doi:10.1016/j.bbmt.2019.02.012

11. Kong W, Yao Y, Zhang J, Lu C, Ding Y, Meng Y. Update of treatment for mucopolysaccharidosis type III (sanfilippo syndrome). Eur J Pharmacol. 2020;888:173562. doi:10.1016/j.ejphar.2020.173562

12. Heldermon CD, Qin EY, Ohlemiller KK, et al. Disease correction by combined neonatal intracranial AAV and systemic lentiviral gene therapy in Sanfilippo Syndrome type B mice. Gene Ther. 2013;20(9):913–921. doi:10.1038/gt.2013.14

13. Fu H, DiRosario J, Kang L, Muenzer J, McCarty DM. Restoration of central nervous system alpha-N-acetylglucosaminidase activity and therapeutic benefits in mucopolysaccharidosis IIIB mice by a single intracisternal recombinant adeno-associated viral type 2 vector delivery. J Gene Med. 2010;12(7):624–633. doi:10.1002/jgm.1480

14. Abeona Therapeutics Receives FDA Fast Track Designation for ABO-101 for Treatment of Sanfilippo Syndrome Type B (MPS IIIB). https://investors.abeonatherapeutics.com/news-events/press-releases/detail/148/abeona-therapeutics-receives-fda-fast-track-designation-for-abo-101-for-treatment-of-sanfilippo-syndrome-type-b-mps-iiib. Accessed on May 16, 2026

15. Gene Transfer Clinical Trial for Mucopolysaccharidosis (MPS) IIIB (MPSIIIB). https://clinicaltrials.gov/study/NCT03315182?viewType=Card&term=Abeona%20Therapeutics&rank=3. Accessed on May 16, 2026

16. Intracerebral Gene Therapy in Children With Sanfilippo Type B Syndrome. https://clinicaltrials.gov/study/NCT03300453?viewType=Card&term=UniQure%20Biopharma&rank=3. Accessed on May 16, 2026

17. Tardieu M, Zérah M, Gougeon ML, et al. Intracerebral gene therapy in children with mucopolysaccharidosis type IIIB syndrome: an uncontrolled phase 1/2 clinical trial. Lancet Neurol. 2017;16(9):712–720. doi:10.1016/S1474-4422(17)30169-2

18. Ma X, Zhuang L, Ma W, et al. AAV9-Mediated Gene Therapy for Infantile-Onset Pompe’s Disease. N Engl J Med. 2025;392(24):2438–2446. doi:10.1056/NEJMoa2407766

19. Whiteman P, Young E. The laboratory diagnosis of Sanfilippo disease. Clin Chim Acta. 1977;76(1):139–147. doi:10.1016/0009-8981(77)90126-7

20. Chuang CK, Lin HY, Wang TJ, Tsai CC, Liu HL, Lin SP. A modified liquid chromatography/tandem mass spectrometry method for predominant disaccharide units of urinary glycosaminoglycans in patients with mucopolysaccharidoses. Orphanet J Rare Dis. 2014;9:135. Published 2014 Sep 2. doi:10.1186/s13023-014-0135-3

21. Gougeon ML, Poirier-Beaudouin B, Ausseil J, et al. Cell-Mediated Immunity to NAGLU Transgene Following Intracerebral Gene Therapy in Children With Mucopolysaccharidosis Type IIIB Syndrome. Front Immunol. 2021;12:655478. Published 2021 May 10. doi:10.3389/fimmu.2021.655478

22. Muenzer J, Ho C, Lau H, et al. Community consensus for Heparan sulfate as a biomarker to support accelerated approval in Neuronopathic Mucopolysaccharidoses. Mol Genet Metab. 2024;142(4):108535. doi:10.1016/j.ymgme.2024.108535

23. MUCOPOLYSACCHARIDOSIS, TYPE IIIB; MPS3B. https://omim.org/entry/252920. Accessed on May 16, 2026.

